# High Prevalence of Malignancy in Obstructive Jaundice: A Cross-Sectional Study from Dongola, Sudan

**DOI:** 10.64898/2026.07.23.26358781

**Authors:** Mohammed Said, Marwa Osman

## Abstract

**Background:** Obstructive jaundice is a clinical condition caused by obstruction of the bile flow system and manifests as discolouration of the eyes and skin. The prevalence of malignant jaundice in nearby African nations is unacceptably high. Furthermore, there is local suspicion that gold mining activities are behind the high number of cancer patients in the northern region of Sudan. This study aims to identify the prevalence of malignancy among obstructive jaundice patients in the study area.

**Methods:** We performed a hospital-based cross-sectional review of 60 consecutive patients diagnosed with obstructive jaundice at three major hospitals in Dongola between January and July 2024. Malignancy was defined by imaging and histopathology. Data were extracted from hospital records and supplemented by telephone interviews when records were incomplete. Descriptive statistics, bivariate tests, and multivariable logistic regression were used to compare malignant and benign cases; significance was set at p < 0.05. Analyses were performed using SPSS v27.

**Results:** Malignancy accounted for 31/60 (51.7%) of obstructive jaundice cases. The most frequent malignant diagnoses were hepatocellular carcinoma (9/31, 29.0%) and carcinoma of the pancreatic head (8/31, 25.8%). Malignant cases more often reported weight loss, vague abdominal pain, dark urine, and pruritus, whereas right upper quadrant pain was more frequent in patients with benign disease. Mean serum bilirubin was significantly higher in malignant versus benign cases (11.2 mg/dL vs 5.0 mg/dL). In multivariable analysis (complete cases, events = 22), age and serum bilirubin remained independent predictors of malignancy (age aOR 1.47 per 10 years, 95% CI 1.07–2.01; bilirubin aOR 2.03 per 5 mg/dL, 95% CI 1.86–2.45). Metastasis showed a strong trend but did not reach significance. Gold mining exposure was significantly associated with malignancy in univariate analysis but lost significance when undergoing multivariable regression testing. Smoking and sex were not significant.

**Conclusion:** In this hospital series from Dongola, over half of obstructive jaundice presentations were due to malignancy, with hepatocellular carcinoma and pancreatic head carcinoma being the most common. High bilirubin levels and older age were independent predictors of malignancy. Residence near gold mining areas was associated with malignancy in descriptive analyses but did not remain significant after adjustment; this finding is hypothesis-generating and requires environmental and epidemiologic follow-up.

## Introduction

Jaundice is a common complaint in hospitals and primary healthcare facilities. It is defined as the yellow discolouration of the sclera and skin caused by an increased serum bilirubin (a by-product of haemoglobin metabolism). This elevation in bilirubin occurs due to overproduction (from red blood cell haemolysis) or bile retention in the liver or biliary tree. The aetiology of jaundice is classified as prehepatic, hepatic, and post-hepatic [1].

Post-hepatic (obstructive) jaundice results from any resistance to bile flow, whether indigenous to the biliary tree or from mass lesions compressing the pathways through which bile passes to reach the intestines. The causes of obstructive jaundice can be divided into benign causes (such as stones, inflammation, surgery-induced strictures, etc.) and malignant causes (tumours of the liver, biliary system, or nearby organs) [1–3].

The most common tumours causing obstructive jaundice are pancreatic head tumours, gallbladder carcinoma, cholangiocarcinoma, and periampullary carcinoma [2]. Pancreatic carcinoma is one of the leading causes of terminal tumours, with a median survival time of 11 months (8.1% 5-year survival rate) for unresected tumours, which increases to a median survival time of 25 months (23.6% 5-year survival rate) after resection [3]. Cholangiocarcinoma also has a grim prognosis, with a median survival time of 10 months that rises to 33 months following successful surgical resection [4]. Gallbladder carcinoma is even worse, with a median survival time of only 5 months (4.5% 3-year survival rate that drops to 2.9% if obstruction causing jaundice is present). Chemotherapy increases the 3-year survival rate to 35% [5]. Periampullary carcinoma has a median survival time of 26 months (24.3% 5-year survival rate), with treatment increasing the 5-year survival rate to 31.6% [6].

Many studies have investigated the causes of obstructive jaundice to determine how often malignancies cause cholestatic jaundice. Some studies show that 56% of study populations who presented with obstructive jaundice were found to have a tumour [7]. Other studies demonstrate a range between 40% and 70% of obstructive jaundice cases being caused by malignancies.

Typically, patients with obstructive jaundice present with itching due to bile salt deposition under the skin, in addition to indigestion of fatty foods due to the entrapment of bile salts. The inability of bile to reach the intestines also retards the formation of the pigments (stercobilin and urobilinogen) that give stool and urine their usual colours, leading to pale stool and dark urine. In addition, pain after meals, resulting from the gallbladder attempting to secrete bile against resistance, causes a degree of malnutrition that manifests as weight loss. On the other hand, patients with cancer may present with cachexia, which is weight loss due to metabolic derangements caused by the tumour or the chronic inflammatory response mounted by the body against the tumour [1].

Sudan’s Northern State is part of the African Sahara, with most of the population concentrated on the banks of the River Nile. It is known as Nubia, “the land of gold”, and has historically attracted gold miners [8]. This activity is mostly carried out without government control, allowing traditional artisanal mining techniques (which heavily depend on mercury) to predominate (Figure 1). Consequently, mercury poisoning cases are frequently encountered in healthcare facilities in the north. Another health risk around gold mining areas is the increased level of radiation, reaching up to about 50% above the global normal range [9]. Despite investigations into the risks of radiation around gold mining sites in different parts of Sudan, the annual gonadal radiation dose has been found to be increased around gold mines. However, the specific element causing the radiation remains unidentified [9]. Moreover, Kordofan is another region in Sudan with a high incidence of cancer (namely nasopharyngeal carcinoma), likely linked to increased radiation caused by uranium in the soil [10], in addition to the presence of multiple gold mining zones in the Nuba Mountains region.

**Figure 1.**
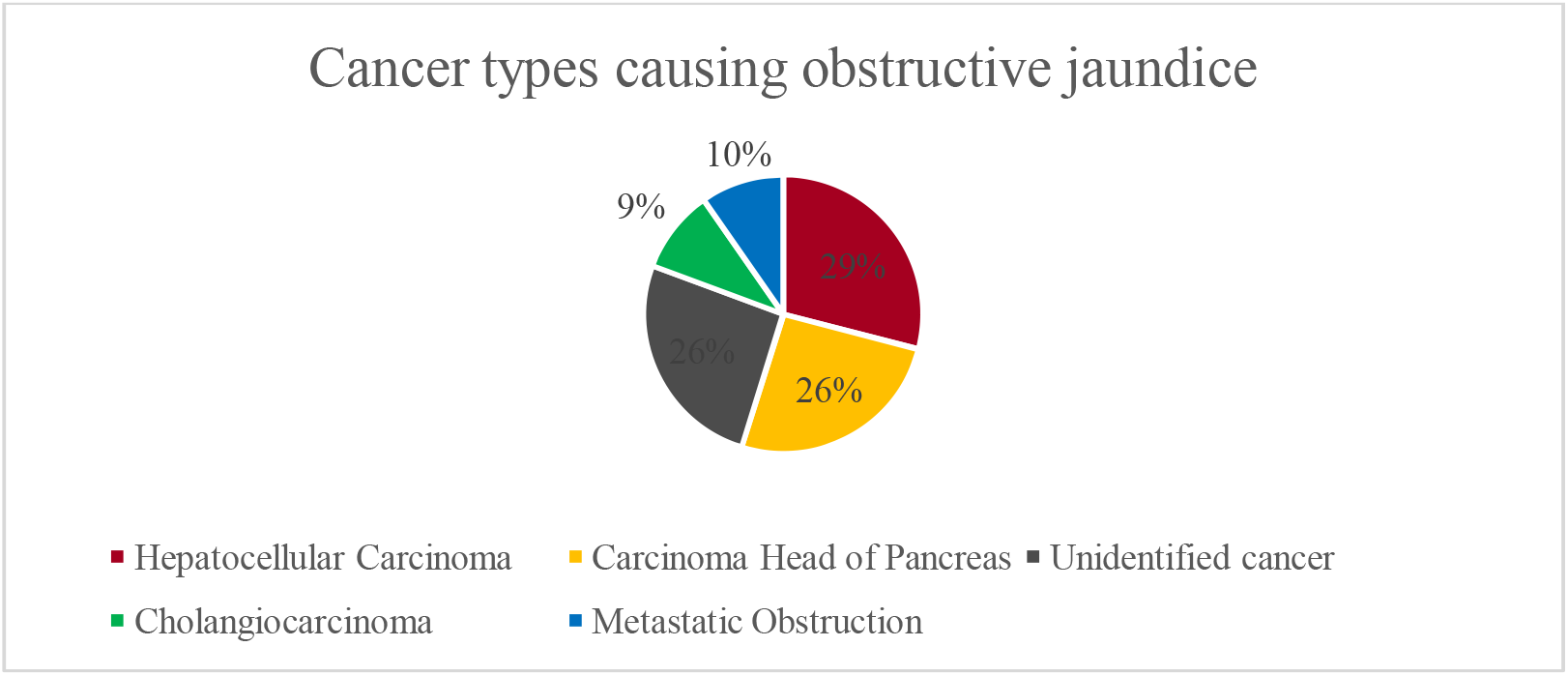
Percentages of cancer types causing obstructive jaundice (n=31).

**Figure 2.**
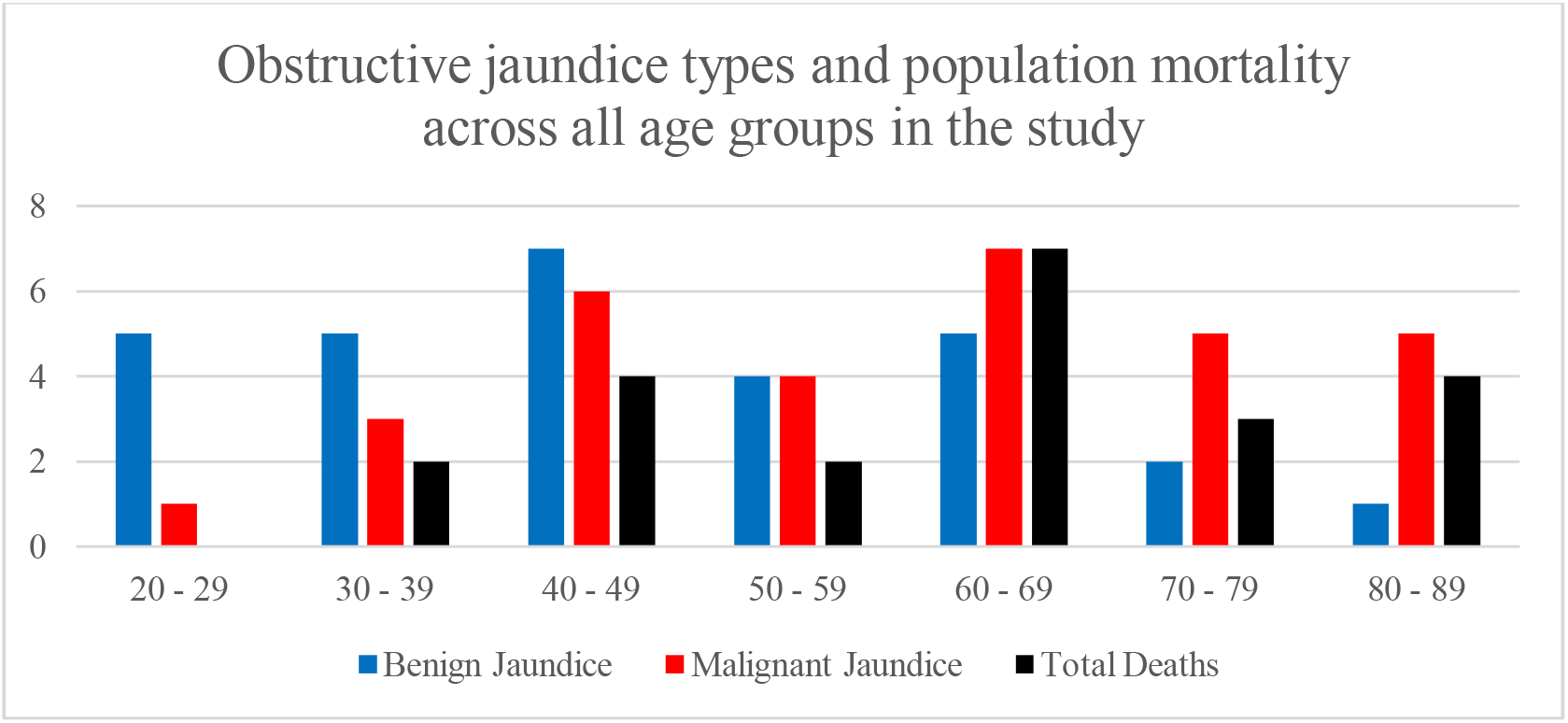
Distribution of Obstructive Jaundice types and mortality amongst the age groups of the study population.

Popular theories among Northern State citizens point to certain practices in the food industry, incidences of radioactive waste burial in the desert, and the widespread extra-governmental gold mining in the north as the reasons behind the observed increased incidence of tumours in general, and gastrointestinal tumours (pancreas, gallbladder, liver) in particular. Within these informal narratives, all of the above are based on raw observations that have not been formally investigated. This inevitably leads to inefficient allocation of limited resources in an already burdened healthcare system. More importantly, the most common malignant tumours causing obstructive jaundice are notorious for their aggressive nature, with time being the determining factor in patient survivability. Time can be lost when a serious diagnosis, such as a pancreatic tumour, is treated at the same pace as a gallbladder stone.

With the variation in the incidence of malignancies causing obstructive jaundice reported across different studies internationally, the need arises to investigate how often a tumour is responsible for jaundice. This study aims to determine the frequency and highlight the presence of any risk factors for cancer among patients presenting with obstructive jaundice in Dongola’s three largest hospitals. This knowledge is essential to improve the efficiency of clinical work-up and management protocols.

## Methods

### Study design and setting

A hospital-based cross-sectional review of patients presenting with obstructive jaundice at AlShamal Specialised Hospital, Dongola Specialised Hospital, and Dongola Police Hospital between January 1^st^ and July 31^st^ 2024.

### Participants and case definition

Consecutive patients with a clinical and/or radiological diagnosis of obstructive (post-hepatic) jaundice were eligible. Patients whose diagnosis of obstructive jaundice was not confirmed or was later revised to a non-obstructive cause were excluded. Malignant versus benign etiology was determined by imaging and histopathology when available, or by the treating clinician’s final diagnosis recorded in the hospital file.

### Data collection

Data were extracted from hospital records using a pretested form and supplemented by telephone interviews when records were incomplete. The collection process extended from August 5^th^ to the 16^th^, 2024. The identifying information were immediately coded, and patients’ phone numbers were deleted after the end of data collection phase. Variables included demographics, presenting symptoms (pain pattern, weight loss, stool and urine color, pruritus), social exposures (smoking, oral tobacco, occupational history including gold mining work, residence near gold mining areas), laboratory results (serum bilirubin when available), imaging and diagnostic findings, and outcome (alive or deceased at the time of data collection).

### Statistical analysis

Continuous variables are presented as mean ± SD or median (IQR), as appropriate; categorical variables as counts and percentages. Bivariate comparisons used chi-square or Fisher’s exact tests for categorical variables and t-tests or Mann–Whitney U tests for continuous variables. Correlations are reported where relevant. Multivariable logistic regression was used to estimate adjusted associations between malignancy and selected predictors (age, sex, serum bilirubin, gold mining exposure, smoking). Complete case analysis was used for multivariable models; model diagnostics and multicollinearity checks were performed. However, due to the small sample size, the results of the model become unstable. Statistical significance was set at p < 0.05. Analyses were performed using SPSS v27.

### Ethical consideration

This study was conducted in accordance with the Declaration of Helsinki and was formally approved by the University of Khartoum, Faculty of Medicine Institutional Review Board / Ethics Committee (Protocol ID: COMMED 2024-95-16). Verbal informed consent was obtained from participants for telephone follow-up and supplementary data collection; the ethics committees approved the use of routinely collected clinical data with verbal consent.

## Results

### Baseline characteristics

Sixty patients were included in the study, of whom 31 (51.7%) had malignant obstructive jaundice and 29 (48.3%) had benign causes. The mean age of the cohort was 53 years (SD = 17.7, range 22–85). Females comprised 56.7% (n = 34). Malignancy was more common in older age groups, particularly 60–69 years, whereas benign disease predominated among patients younger than 50 years.

### Malignant obstructive jaundice

Among malignant cases, hepatocellular carcinoma was the most frequent diagnosis (29.0%), followed by carcinoma of the pancreatic head (25.8%), cholangiocarcinoma (9.7%), and metastatic obstruction (9.7%). Eight patients (25.8%) died before a definitive malignant subtype could be identified. Benign causes included gallstones (41.4%), cholecystitis (37.9%), cholangitis (6.9%), and isolated cases of surgical stricture, pancreatitis, hepatic fibrosis, and cirrhosis.

### Age and gender factors in Malignant obstructive jaundice

Two-thirds of malignant patients were above 50 years (66.7%), with a mean age of 58 years (SD = 17.2), compared to 46 years (SD = 16.6) in benign cases. Age correlated positively with malignancy (Pearson’s r = 0.345, p = 0.007). Logistic regression confirmed age as an independent predictor (aOR 1.47 per 10 years, 95% CI 1.07–2.01, p = 0.018). Gender was not significantly associated with malignancy (p = 0.065), although males were proportionally more affected (65.4% vs 41.2%).

### Disease outcome

The study found a significant association between disease outcome and type of obstructive jaundice. 54.8% of malignant obstructive jaundice patients (n = 17) passed away during or before data collection, compared to 17.2% of benign obstructive jaundice patients (n = 5), who died due to other illnesses. A fatal disease outcome was found to be related to malignant obstructive jaundice (p-value = 0.038).

### Relationship between serum bilirubin and type of obstructive jaundice

Serum bilirubin was measured in 47 patients. Malignant cases had significantly higher levels (mean 11.2 mg/dL, SD = 10.2, range 3.1–41.7) compared to benign cases (mean 5.0 mg/dL, SD = 2.2, range 2.9–13.7; Mann–Whitney U, p = 0.005). Bilirubin correlated with malignancy (Pearson’s r = 0.41, p < 0.001) and remained an independent predictor in regression (aOR 2.03 per 5 mg/dL, 95% CI 1.86–2.45, p < 0.001).

### Symptoms of malignant obstructive jaundice

All patients presented with jaundice. Symptom profiles differed markedly between groups:

- Cachexia: Malignant 90.3% vs benign 17.2% (Pearson’s r = 0.734, p < 0.001).
- Abdominal pain (vague): Malignant 80.6% vs benign 31.0% (r = 0.500, p < 0.001).
- RUQ pain: Benign 69.0% vs malignant 19.3% (p < 0.0001); logistic regression confirmed an inverse association (aOR 0.13, 95% CI 0.06–0.24, p < 0.001).
- Dark urine: Malignant 71.0% vs benign 24.1% (r = 0.468, p < 0.001).
- Itching: Malignant 32.3% vs benign 10.3% (r = 0.266, p = 0.04).
- Other symptoms (nausea, vomiting, pale stool, fever/rigours): No significant differences (all p > 0.1).

A particular relationship was found between individual cancers and certain symptoms, as shown in Table 2 below. A noteworthy finding is that only 6 out of 31 (19.4%) cancer patients had painless jaundice.

**Table 1:** Age and gender characteristics of the participants n=60.

| Age group | Females | Males | Total |
| --- | --- | --- | --- |
| 20 – 29 | 4 | 2 | 6 |
| 30 – 39 | 3 | 5 | 8 |
| 40 – 49 | 9 | 4 | 13 |
| 50 – 59 | 6 | 2 | 8 |
| 60 – 69 | 7 | 5 | 12 |
| 70 – 79 | 4 | 3 | 7 |
| 80 – 89 | 1 | 5 | 6 |
| Total | 34 | 26 | 60 |

**Table 2:** Symptoms of benign and malignant obstructive jaundice patients (n=60)

| Symptoms | Benign obstructive jaundice patients (N=29) (100%) | Malignant obstructive jaundice patients (N=31) (100%) |
| --- | --- | --- |
| RUQ pain | 20 (69) | 6 (19.3) |
| Shoulder pain | 10 (34.5) | 2 (6.4) |
| Vague abdominal pain | 9 (31) | 25 (80.6)<br>($p < 0.001$) |
| Pale stool | 16 (55.2) | 23 (74.2) |
| Dark urine | 7 (24.1) | 22 (71)<br>($p < 0.001$) |
| Nausea | 15 (51.7) | 18 (58) |
| Vomiting | 13 (44.8) | 18 (58) |
| Fever or rigours | 7 (24.1) | 5 (16.1) |
| Itchiness | 3 (10.3) | 10 (32.3) |
| Cachexia | 5 (17.2) | 28 (90.3)<br>$p < 0.001$ |

### Social factors

- Smoking: Malignant 39% vs benign 17% (p = 0.069; logistic regression aOR 1.07, 95% CI 0.58–1.96, ns).
- Oral tobacco: No significant association (p = 0.36).
- Family history of similar disease: Benign 34% vs malignant 13% (p = 0.051, ns).
- Family history of jaundice: No difference (p = 0.85).
- Gold mining exposure: Malignant 77% vs benign 41% (p = 0.047). Correlation with cholangiocarcinoma was significant (r = 0.358, p = 0.005). However, it did not remain significant in multivariable testing. No association was found in females (p = 0.816).

## Discussion

The primary objective of this study was to calculate the frequency of cancers causing obstructive jaundice presentations at the three main hospitals in the city of Dongola, the capital of the Northern State. Sixty patients were identified with reference to general surgeons, gastroenterologists, and hospital files. Malignancies were found in 51.7% of patients who presented with jaundice that was determined to be obstructive. This percentage is close to the studies conducted in Pakistan, which reported 52% and 57% [11, 12], Nepal at 50.9% [14], the UK at 53.2% [21], and Tanzania, an East African nation, reported a 58.7% incidence of malignancies among obstructive jaundice patients [17]. However, the percentage in this study is unacceptably high, especially with some nearby nations reporting an incidence as low as 5% in southwestern Saudi Arabia [16].

The second goal of this study was to identify the most common cancer type behind obstructive jaundice presentations. The findings agree with international literature that carcinoma of the head of the pancreas is very commonly the reason behind the obstruction to bile flow [2,3,6]. However, hepatocellular carcinoma (HCC) was found to be more common in the study cohort by a narrow margin, despite usually being a rare cause of obstructive jaundice. Although HCC is typically an uncommon cause of obstructive jaundice, its higher frequency in this cohort may reflect local epidemiological factors such as high hepatitis B and C prevalence. However, given the small sample size and diagnostic limitations, this finding should be interpreted cautiously. This finding is different from all mentioned literature, where primary liver cancers are the least likely causes of obstructive jaundice [28,29,30]. In addition to the persistent taboo usage of alcohol, this may subsequently increase the possibility of liver cancers. Rare variants of HCC can present with obstructive jaundice, but this remains uncommon in most series. The cholestatic variant of HCC carries an equally grim prognosis to the other malignant jaundice cancers [26]. However, patients can benefit from surgical management [29].

The third goal of this study was to identify the particular symptoms that accompany obstructive jaundice of malignant origin. The results regarding painless jaundice as an indicator of cancer disagree with common knowledge [2, 23, 27]. Many studies indicate that most cancer patients present with painless jaundice and that around 40% or fewer have pain as a complaint with malignant jaundice [14, 15, 23, 27]. The findings in this study indicate that 19.4% had painless jaundice. The remaining 80.6% of cancer patients reported vague abdominal pain that could not be localised. Upon localisation of pain, the prognosis shifts to a more benign cause (p < 0.001). A similar finding was reported in multiple studies [13, 25, 31], which suggests variability and warrants further investigation.

The study also found a stronger probability of malignancy in jaundice patients who complained of weight loss, dark urine, and itching. This finding agrees with the literature and general knowledge [2,4,5]. The manifestation of dark urine takes longer in comparison to pale stool; therefore, the discolouration of urine is a reliable tell-tale sign of a longer and more insidious pathology. Itching from bile salt deposition under the skin aligned with high serum bilirubin levels [1] (a feature that correlates with malignant obstructive jaundice in this study). Therefore, it can be considered a sign of marked hyperbilirubinemia and a diagnostic indicator of malignancy. On the other hand, patients reporting RUQ abdominal pain or radiation of pain to the shoulder were mostly suffering from a benign condition. This distinction in presentation between benign and malignant obstructive jaundice types is not frequently mentioned in the literature, making confirmation of this finding difficult.

A reliable diagnostic indicator of a malignant cause in the study population was the serum bilirubin level (p = 0.005). This finding aligns with the literature and supports increased clinical suspicion of malignancy, particularly at higher levels [18, 19, 20, 21, 22].

The final goal of this research was to identify risk factors for malignant obstructive jaundice causes as a group. Each of the cancers has its well-outlined risk factors. However, the aim was to investigate the presence of factors causing susceptibility of the hepatobiliary and pancreatic systems to cancer development. The study found two risk factors shared between all cancer types: old age (p = 0.007) and proximity to gold mining regions (p = 0.047). Despite smoking being a known risk factor for carcinoma of the head of the pancreas and cholangiocarcinoma, it was not a significant risk factor among the studied cancers or malignant jaundice. Old age is a well-known factor in the development of malignancies [1]. The findings agree with the literature and current knowledge. The majority of cancer patients across different studies presenting with jaundice are above 60 years of age. This fact seems to have remained constant since the 1980s. The overall incidence of malignant obstructive jaundice among older patients is significantly higher than in younger populations [20].

The relationship between the development of cancers and proximity to gold mining regions is hypothesis-generating. The study found an association between cholangiocarcinoma and inhabitants of places close to gold mines (p = 0.005), but adjustment for other factors rendered it non-significant independently. These findings may be a consequence of the effects artisanal gold mining has on the environment [32]. Gold mining is a major source of toxic heavy metal deposition in soil, water, and eventually the living population. A study conducted by Ahmed Eltohami indicated that regions surrounding gold mines have health hazards [24] and an increased annual gonadal dose equivalent measured in people living nearby [9]. The annual gonadal dose equivalent indicates the quantity of radiation absorbed into living tissue. This particular study excluded the presence of uranium or other major radioactive elements from an eastern region in Sudan that surrounds a gold mine, but still found an increased level of radiation that could be linked to gold mining activity and the metals used in the process [9]. The findings related to gold mining in this study should be interpreted with caution due to the observational design and limited sample size.

## Limitations to this study

The design and sample size—a single-city, hospital-based, cross-sectional design with a modest sample (n = 60)—limit generalizability and causal inference.

- Serum bilirubin was not measured in all patients (available for 47/60); complete-case multivariable models excluded 13 records. Denominators are reported for each analysis.
- Although malignancy was defined by imaging and histopathology when available, some patients died before a definitive malignant subtype could be identified (8/31).
- Gold mining exposure was self-reported and approximated by residence; no direct environmental or biomarker measurements were available.
- Multivariable models had limited events per predictor; some adjusted estimates are imprecise. Sensitivity analyses (penalized regression or multiple imputation) would strengthen inference.

## Conclusion

Results from this study concluded that half of the patients presenting with obstructive jaundice have cancer underlying the presentation. There is no significant difference between genders in susceptibility. The most common cancer type underlying bile flow obstruction in our cohort is hepatocellular carcinoma, followed closely by carcinoma of the head of the pancreas. The symptoms associated with malignant obstructive jaundice are weight loss, dark urine, itching, and abdominal pain that cannot be localized. A high bilirubin level is among the strong indicators of malignancy. Age is a shared risk factor for all causes of malignant jaundice, which is supported by studies. Proximity to gold mining areas or working in gold mining may be associated with an increased risk of cancer development, particularly in males. Most malignant obstructive jaundice patients present late, in the terminal stage, which explains the high mortality rates, in addition to the grim survival chances associated with the studied cancers.

## Recommendations

The lethality of the condition and its implications for quality of life make further investigations mandatory. Larger sample sizes and longer-duration studies (prospective cohorts) must be conducted in future research to allow greater understanding of all factors at play.

Moreover, the possibility of artisanal gold mining being a health hazard, and a probable cause of carcinogenesis, warrants further research. Adding cancer to the health risks of local, unsanctioned artisanal gold mining will hopefully encourage more regulations in favor of public health.

## Data Availability

All data produced in the present study are available upon reasonable request to the authors

## Abbreviations

CA: Carcinoma
HCC: Hepatocellular Carcinoma
RUQ: Right Upper Quadrant

